# Parametric Models for Predicting Gram-negative Antibiotic Susceptibility Phenotype: A Brief Report

**DOI:** 10.64898/2026.10.01.26363073

**Authors:** Akhil Garg, Christopher Graham, Nick Daneman, Derek MacFadden

## Abstract

Predicted probabilities of antibiotic susceptibility to known or suspected pathogens can be used to support antibiotic selection. Patient demographic, clinical, and prior microbiologic history can be used to generate location-specific prediction models. For three Canadian tertiary care institutions we fit and evaluated models for predicting Gram-negative antibiotic susceptibilities to common antimicrobial agents based on previously established model structures. A web-based application was developed to permit visualization of antibiotic susceptibility predictions and support antibiotic decision-making.

## BACKGROUND

A provider’s empiric antibiotic selection in suspected infection should be tailored to the individual’s risk of antibiotic resistance^1–4^. Treatment guidelines have historically recommended simple heuristics to help inform providers of a patient’s risk for a resistant pathogen^4,5^, but these lack the ability to provide more patient-specific risk across a variety of epidemiologic risk factors. Traditional antibiograms have also been incorporated into guidelines, but these represent an average risk across a broad range of patients (e.g. an entire hospital, ward, or culture type) and are not informed by an individual’s specific risk factors^6^. Previous work has evaluated acceptable treatment probability thresholds for clinicians when treating infection of differing severity, in order to operationalize individual probability of susceptibility estimates, with 80% and 90% probabilities of susceptibility found to be acceptable for non-severe and severe infection^3^. Moreover, treatment guidelines are incorporating antibiotic susceptibility/resistance likelihood thresholds into therapy selection recommendations^5,7^. However, easy and generalizable approaches to generate integrated and personalized resistance risk estimates are needed.

Simple, parametric models, based on common but important epidemiologic risk factors for resistance (e.g. age, sex, location of acquisition, prior antibiotic exposure, prior microbiologic history) offer the potential for individualized risk assessment^2,8,9^.

We have previously demonstrated the utility of parametric models, based on epidemiologic and clinical variables, for predicting antibiotic susceptibility across Gram-negative pathogens^2,8,9^.

In this brief report we sought to: (1) fit and evaluate prediction models at three separate tertiary care institutions based on previously demonstrated model structures and approaches; and (2) develop and share a web-based application for visualization of antibiotic susceptibility predictions using model covariates.

## METHODS

### Study Design

We performed a retrospective observational study of hospitalized patients with suspected infection with confirmed Gram-negative organisms across three tertiary care health centers in Canada (denoted hereafter as Sites A, B, and C). Institution specific prediction models were developed and evaluated for each site. A web-based application was developed to support visualization of antibiotic susceptibility prediction data. Research ethics board approvals, including waiver of individual consent for data collection, were obtained for this study.

### Population/Data

We identified Gram-negative isolates from episodes of suspected infection in admitted adult patients at each participating tertiary care institution covering a span of 2-4 years between 2016 to 2023. These data were obtained from institutional electronic medical record systems and/or clinical stewardship databases. Suspected infection was defined as both initiation of broad-spectrum antibiotic therapy (3^rd^ generation cephalosporins, fluoroquinolones, carbapenems, and/or aminoglycosides) and concurrent isolation of a Gram-negative bacteria (monomicrobial) from clinical culture (within 24 hours). Gram-negative susceptibility data were linked with demographic and clinical data. Cultures from repeat episodes within 24 hours were excluded, along with polymicrobial cultures, or cultures without any susceptibility data.

### Outcomes

The primary outcome of interest was phenotypic Gram-negative antibiotic susceptibility, across five major antibiotics (Meropenem, Piperacillin/Tazobactam, Ceftazidime, Ceftriaxone, and Ciprofloxacin). Antibiotic susceptibility was collected from institutional medical record data and reflect clinical microbiologic laboratory testing results at respective sites. All sites used MALDI-TOF for organism identification, and VITEK or BD Phoenix-based antibiotic susceptibility testing according to CLSI standards. Intermediate and resistant phenotypes were both considered ‘non-susceptible’ (hereafter termed resistant) phenotypes. Site-specific (rules-based) susceptibility imputation was performed based on previously described methods ^10,11^. Any remaining missing susceptibility values were assumed to be resistant during data analyses.

### Covariates

Predictors for the parametric models were taken from previously described model structures, and these included age category, sex, acquisition location (community vs. hospital non-critical care unit vs. critical care unit), recent hospitalization, admitting service, prior antimicrobial exposures by class, prior culture results, and prior ESBL history^2,8^. This parsimonious set of predictors (described in detail in Supplementary Table 1) were selected *a priori* based on the existing clinical data supporting their use in these model structures for these purposes, and to reduce risk of over-fitting (by not including a data-driven covariate selection step). Covariate harmonization was performed, but there remained minor site-specific differences in definitions, and these are summarized in Supplementary Table 1.

### Data Analyses

Logistic regression models were fit for each antibiotic susceptibility phenotype (*i*.*e*. meropenem, piperacillin-tazobactam, ceftazidime, ceftriaxone, and ciprofloxacin) for each site and respective dataset, in a similar fashion as previously described^2^. All described predictors were used for each model – there were no parameter selection/removal stages. Model coefficients are reported for each antibiotic phenotype and site. Area under the receiver operating characteristics curve (AUC) statistics were generated for each site and antibiotic susceptibility phenotype. To better evaluate discrimination while accounting for potential overfitting, time, and clustering effects, AUCs were also estimated using temporally split derivation (75-80%) and validation (25-20%) subsets. Calibration of the models was confirmed by binning predicted marginal probabilities into deciles and comparing with susceptibility outcome proportion. Calibration was evaluated and reviewed for each scenario, with a focus on relevant clinical treatment thresholds of 80% and 90% probability of susceptibility, which are reported for the full models by site and phenotype. Data analyses were performed using Python v3.13.3.

### Web-based Application Development

To facilitate visualization of antibiotic susceptibility predictions, we developed a web-based app using the Shiny platform^12^. This app was developed to allow toggling between multiple institutions (with differing predictor/covariate definitions – that show as smart tool hover-overs) and generate real-time simultaneous visualization of predicted probability of susceptibility across different antibiotic phenotypes. Relevant clinical thresholds of 80% and 90% were included as markers in the visualization, though this can be adjusted. The predictor values can be varied in real-time to modify predicted probabilities of susceptibility. This application was developed using Python v3.13.3, and the code for this app is available at http://github.com/A-Garg/IDEAS-Demonstration/.

### Data Availability

Data used in the study are available upon reasonable request to the corresponding author.

## RESULTS

Across the three sites, there were 11,402 Gram-negative cultures identified, including 2,948 for Site A, 2,985 for Site B, and 5,469 for Site C. Across all sites, just over half of cultures (52%) were in female individuals, and the most common age category was 75-80 years (13%). Most were collected in the context of community acquisition (64%), with 28% involving recent hospitalization and 77% occurring during medical admissions. Prior antibiotic exposure was common, ranging from 10-33% depending upon class. Prior ESBL isolates were present in 5%, and 20% had prior Gram-negative susceptibility results available. Less than 25% of cultures across all sites represented a repeated test for a given patient. Baseline antibiotic susceptibility prevalence varied by class and site and is summarized in Table 1. Model parameters, by antibiotic phenotype and site, are summarized in Supplemental Table 2. AUC values were similar between the temporal split sample and full model estimation (Table 2), with medians of 0.72 (IQR 0.7-0.75) and 0.72 (IQR 0.71-0.76) respectively. Calibration was consistent across the models, with a median value of 0.85 (IQR 0.84-0.85) for the 80-90% threshold, and a median value of 0.95 (IQR 0.92-0.98) for the 90-100% threshold. Calibration remained robust across changes in beta-lactam susceptibility data resulting over time.

**Table 1.** Baseline antibiotic phenotypic susceptibility prevalence by site.

| Site | Prevalence of Antibiotic Phenotypic Susceptibility |  |  |  |  |
| --- | --- | --- | --- | --- | --- |
|  | Ciprofloxacin | Ceftriaxone | Ceftazidime | Piperacillin/Tazobactam | Meropenem |
| Site A<br>(n=2948) | 78 | 70 | 81 | 76 | 95 |
| Site B<br>(n=2985) | 77 | 64 | 75 | 83 | 98 |
| Site C<br>(n=5469) | 75 | 58 | 71 | 70 | 95 |

**Table 2.** Model calibration and sample discrimination (AUC) values, by antibiotic susceptibility phenotype, and site.

| Site | Antibiotic Phenotype | AUC* | Calibration at 80-90% | Calibration at 90-100% |
| --- | --- | --- | --- | --- |
| Site A | Ciprofloxacin | 0.71/0.69 | 0.85 | 0.92 |
|  | Ceftriaxone | 0.75/0.74 | 0.85 | 1 |
|  | Ceftazidime | 0.69/0.70 | 0.87 | 0.88 |
|  | Piperacillin-Tazobactam | 0.72/0.71 | 0.85 | 1 |
|  | Meropenem | 0.86/0.84 | 0.85 | 0.97 |
| Site B | Ciprofloxacin | 0.67/0.66 | 0.84 | 0.94 |
|  | Ceftriaxone | 0.75/0.75 | 0.84 | 0.97 |
|  | Ceftazidime | 0.7/0.72 | 0.86 | 0.95 |
|  | Piperacillin-Tazobactam | 0.69/0.71 | 0.86 | 0.92 |
|  | Meropenem | 0.75/0.85 | 0.84 | 0.99 |
| Site C | Ciprofloxacin | 0.67/0.68 | 0.84 | 0.93 |
|  | Ceftriaxone | 0.77/0.77 | 0.85 | 0.86 |
|  | Ceftazidime | 0.72/0.71 | 0.84 | 0.94 |
|  | Piperacillin-Tazobactam | 0.73/0.72 | 0.84 | 0.96 |
|  | Meropenem | 0.86/0.85 | 0.84 | 0.98 |

\*Temporally Split Sample AUC/AUC.

## DISCUSSION

Antibiotic susceptibility varied by site and antibiotic but was broadly consistent with other published studies evaluating Gram-negative susceptibility^2,13^.

AUCs and calibration were consistent across evaluated sites and antibiotics with previously published literature for predicting Gram-negative susceptibility from common, readily available, epidemiologic predictors^2,14^.

One inherent limitation of these predication models is that they are institution specific (i.e. only valid for the evaluated hospital population and associated bacterial ecology) and are thus not expected to be generalizable to other health centers. However, these model structures generate consistent performance, are straightforward to fit, and are suitable for tailored, location-specific antibiotic phenotype predictions, enabled by increasingly utilizable microbiologic and EMR data.

While these data were predominantly composed of single culture result observations, additional cultures meeting the eligibility criteria for an individual were included (enabling predictions for later hospital or healthcare exposed episodes). Measures of model discrimination may be influenced by presence of repeat observations, but this is unlikely to impact our findings given: (1) the pre-specified model structure helps mitigate overfitting; (2) additional observations for any individual represented a minority of data points; and (3) temporally aligned split evaluation (which helps to keep clustered observations in time for individuals in the same subset) was consistent with the overall estimate. Moreover, a dataset sensitivity analysis restricting the validation subset to unique observations also yielded comparable results.

A web-based Shiny application was developed to support the visualization of antibiotic susceptibility predictions, and a demonstration of the web interface is available at http://akhilgarg.shinyapps.io/dummymodel/.

In summary, simple institution-specific parametric models based off of routine epidemiologic and clinical predictors can be used to predict probability of antibiotic susceptibility in Gram-negative isolates and can be visualized using web-based tools.

## Notes

### Competing Interest Statement

The authors have declared no competing interest.

### Author Declarations

Ethics committees/IRBs of The Ottawa Hospital, Sunnybrook Health Sciences, and Trillium Health Partners, all gave ethical approval for this work, including waivers of consent for data collection.

