## Supplemental Material for "Parametric Models for Predicting Gram-negative Antibiotic Susceptibility Phenotype: A Brief Report"

### Supplementary Table 1. Covariate definitions.

| Covariate | Definition* |
|---|---|
| Age | Age category at time of index culture collection. |
| Sex | (i) Sex from index encounter;<br>(ii) Sex at time of index culture collection. |
| Clinical ESBL | Clinical culture result in prior 3-365 days containing 'ESBL' or 'Extended' strings. |
| Recent Hospitalization | (i) Admission within 90 days prior to index culture collection date;<br>(ii) Separate hospitalization in 90 days prior to present admission date. |
| ICU Exposure | (i) ICU admission within 90 days prior to index culture collection date;<br>(ii) Admitted to ICU at any point in the last 90 days. |
| Prior Gram-negative resistance | Phenotypic susceptibility (susceptible, non-susceptible) for the most recent Gram-negative in the 3-90 days prior to index culture collection date. Absence of prior isolate or susceptibility result used as reference category. |
| Prior antibiotic use by class | Antibiotic (by class) taken within 2-90 days prior to index culture collection date. |
| Admission type | Admitting hospital service for index inpatient admission (surgical versus medical). |
| Acquisition | Community = Index culture collected within first 48 of admission or while in ED prior to admission. Hospital = culture collection >=48 hours after admission. ICU = culture collected >=48 hours after ICU admission. |

*Where variations could differ by site, multiple definitions are provided via (i) and (ii).

**Supplementary Table 2. Model parameters by site and antibiotic susceptibility phenotype.**

| Site | Model Parameter | Meropenem | Pip/Tazo | Ceftazidime | Ceftriaxone | Ciprofloxacin |
|---|---|---|---|---|---|---|
| **A** | Age | -0.02 | -0.01 | -0.01 | -0.01 | 0.00 |
| | Sex | 0.2 | 0.05 | -0.07 | 0.1 | 0.4 |
| | Clinical ESBL | 0.2 | 1.2 | 1.4 | 1.1 | 1.1 |
| | Recent Hospitalization | -0.6 | -0.2 | -0.07 | -0.1 | -0.1 |
| | Prior GN Isolate with Resistant Phenotype | 2.6 | 1.5 | 1.5 | 1.7 | 1.8 |
| | Prior GN Isolate with Susceptible Phenotype | 0.1 | -0.1 | -0.2 | -0.4 | -0.3 |
| | Prior Antibiotic Use (Same class) | 1.3 | 0.4 | 0.3 | 0.6 | 1.1 |
| | Prior Antibiotic Use (Different class) | 0.3 | 0.5 | 0.3 | 0.4 | -0.4 |
| | Admission Type | -0.2 | 0.1 | 0.2 | 0.1 | -0.2 |
| | Acquisition Hospital | -0.2 | -0.05 | 0.01 | 0.1 | -0.1 |
| | Acquisition ICU | 0.4 | 0.02 | 0.2 | 0.5 | -0.3 |
| **B** | Age | -0.01 | -0.01 | -0.01 | -0.01 | -0.00 |
| | Sex | 0.5 | 0.3 | 0.5 | 0.6 | 0.2 |
| | Clinical ESBL | -0.4 | 1.8 | 1.6 | 1.5 | 1.5 |
| | Recent Hospitalization | -0.1 | -0.2 | -0.2 | -0.2 | -0.2 |
| | Prior GN Isolate with Resistant Phenotype | 3.0 | 1.1 | 1.4 | 1.3 | 1.5 |
| | Prior GN Isolate with Susceptible Phenotype | 0.0 | -1.3 | -1.4 | -1.4 | -0.7 |
| | Prior Antibiotic Use (Same class) | 2.0 | 0.2 | 0.7 | 1.0 | 1.00 |
| | Prior Antibiotic Use (Different class) | 0.6 | 0.6 | 0.4 | 0.6 | 0.3 |
| | Admission Type | 0.3 | -0.3 | -0.1 | -0.1 | -0.4 |
| | Acquisition Hospital | 0.2 | 0.2 | 0.2 | 0.2 | -0.4 |
| | Acquisition ICU | 0.6 | 0.2 | 0.3 | 0.5 | -0.6 |
| **C** | Age | -0.01 | -0.01 | -0.00 | -0.01 | -0.01 |
| | Sex | 0.3 | 0.3 | 0.3 | 0.5 | 0.1 |
| | Clinical ESBL | -0.1 | 1.5 | 1.6 | 1.6 | 1.3 |
| | Recent Hospitalization | 0.02 | -0.1 | -0.05 | -0.1 | -0.3 |
| | Prior GN Isolate with Resistant Phenotype | 2.4 | 1.2 | 1.1 | 1.4 | 1.5 |
| | Prior GN Isolate with Susceptible Phenotype | -0.2 | -0.8 | -0.8 | -0.9 | -0.7 |
| | Prior Antibiotic Use (Same class) | 1.3 | 0.1 | 0.5 | 0.9 | 1.2 |
| | Prior Antibiotic Use (Different class) | 0.8 | 0.8 | 0.4 | 0.4 | 0.2 |
| | Admission Type | -0.5 | 0.1 | 0.2 | 0.01 | -0.02 |
| | Acquisition Hospital | 1.1 | 0.1 | 0.1 | 0.3 | -0.3 |
| | Acquisition ICU | 1.5 | 0.4 | 0.2 | 0.6 | -0.2 |

*Positive coefficients denote an odds ratio (OR) >1 for resistance to the given antibiotic, and negative coefficients denote an OR <1.
